# The Effect of Intradialytic Exercise on Calcium, Phosphorus and Parathyroid Hormone: A Randomized Controlled Trial

**DOI:** 10.1101/2023.03.20.23287492

**Authors:** Mohammadali Tabibi, Kenneth R Wilund, Nasrin Salimian, Saghar Nikbakht, Mahsa Soleymany, Zahra Roshanaeian, Saghar Ahmadi

## Abstract

**Introduction:** Patients with kidney failure experience derangements of circulating markers of mineral metabolism and dysregulation of skeletal and cardiovascular physiology which results in high mortality rate in these patients. This study aimed to evaluate the effect of intradialytic concurrent exercise on regulation of theses abnormalities related parameters in patients receiving hemodialysis (HD).

**Methods:** In this randomized controlled trial conducted in a HD center in Iran, adult patients receiving chronic HD were randomized to intradialytic exercise (60 minutes) in the second hour of thrice weekly dialysis for 6 months (intervention) or no intradialytic exercise (control). The primary outcomes were serum calcium, serum phosphorous and parathyroid hormone levels. Secondary outcomes were serum alkaline phosphatase and calcium-phosphorous product

**Results:** The study included 44 participants randomized to intervention (n=22) or control (n=22). During the 6-month intervention period, significant between-group changes were observed in all primary and secondary outcomes between the intervention and control groups. The analysis showed a significant decrease in serum levels of phosphorous and parathyroid hormone (P < 0.05). Statistical analyses reveal a significant increase in serum calcium (P < 0.05) as well as a significant decrease in serum phosphorous, parathyroid hormone, alkaline phosphatase and calcium-phosphorous product (P < 0.05).

**Conclusion:** Intradialytic exercise performed for at least 60 minutes during thrice weekly dialysis sessions improves bone mineral metabolism in adult patients receiving HD. Further studies should focus on observing the effect of different types of exercise on bone mineral disorders and all-cause mortality in HD patients.

**Trial registration:** ClinicalTrials.gov Identifier: NCT04916743, Registered on June 8th,2021. Registered trial name: The Effect of Intradialytic Exercise on Calcium, Phosphorous and Parathyroid Hormone: A Randomized Controlled Trial

## Introduction

In recent years, there has been a rapid increase in the prevalence of patients with end stage kidney disease (ESKD) that require kidney replacement therapy, including hemodialysis (HD), peritoneal dialysis (PD) or kidney transplant [1]. ESKD patients, especially those on HD and PD, experience high rates of functional impairment, morbidity, hospitalization, and mortality [2, 3]. These outcomes are an urgent priority for patients, caregivers and healthcare professionals [4, 5]. The primary etiological factors responsible for these outcomes include cardiovascular disorders, muscle atrophy and malnutrition [3]. Cardiovascular disease in ESKD patients is associated with traditional risk factors (diabetes, hypertension, sedentary lifestyle) as well as nontraditional factors (anemia, inflammation, abnormal calcium and phosphorus metabolism and oxidative stress) [6]. High levels of calcium, phosphorus, calcium-phosphorus product and parathyroid hormone are associated with all causes of mortality in dialysis patients [7]. Hyperphosphatemia is also a common complication in ESKD. As renal excretion of phosphorus decreases, this results in increased serum levels of phosphorus and hyperphosphatemia. Calcium and parathyroid hormone are phosphorus-dependent. Hyperphosphatemia alone or in combination with hypercalcemia is associated with increased mortality in hemodialysis patients. Phosphate metabolism is regulated by the interaction of the kidneys, bones, and intestines. This balance is impaired in kidney patients, leading to hyperparathyroidism and vascular calcification [8].

Hyperparathyroidism is one of the aggravating causes of anemia in hemodialysis patients, in addition to resistance to erythropoietin treatment. Defects in calcitriol synthesis, which along with parathyroid hormone stimulates calcium reabsorption, causes hypocalcemia and hyperphosphatemia in dialysis patients. In this condition, there is an increase in parathyroid hormone synthesis to regulate calcium and phosphorus homeostasis, leading to secondary hyperparathyroidism [9-11].

Abnormal levels of parathyroid hormone are associated with disorders of the musculoskeletal system and decreased function, which can lead to increased mortality in these patients. Synthesis and the use of muscular energy are affected by elevated levels of this hormone, which in turn, causes interference in amino acids and protein metabolism in the musculoskeletal system. [12,13]. In CKD patients, especially in chronic (dialysis) patients, the respiratory system is also affected because both uremia and high levels of parathyroid hormone endanger lung function [14]. Therefore, dialysis patients have respiratory disorders and decreased aerobic capacity due to vitamin D deficiency [15] and high levels of parathyroid hormone [14]. Indeed, the functional capacity in dialysis patients has been shown to be about 50% of what it is in healthy populations [16].

The regulation of parathyroid hormone is critical for bone metabolism in healthy individuals as well as dialysis patients [17], and may be influenced by physical activity [18]. Indeed, numerous studies in healthy individuals indicate that exercise plays an important role in the regulation of this hormone [17,19-22]. However, little is known about the role of exercise especially concurrent exercise on parathyroid hormone and calcium and phosphorus metabolism in dialysis patients.

## Methods

### Trial design

This study is an open-label, parallel arm, randomized controlled trial with blinded end-points, which was conducted in a medical center in Iran.

### Participants

Individuals were eligible to participate in the study after meeting all of the following inclusion criteria: 1) age ≥ 18 years; 2) receiving regular HD 3 times a week; 3) on HD for at least 1 year, 4) absence of a history of myocardial infarction within the past 3 months; 5) permission from their physician to participate; and, 6) had capacity to provide informed consent to participate in the study. Individuals were excluded if they met any of the following exclusion criteria: 1) cardiac instability (angina, decompensated congestive heart failure, severe arteriovenous stenosis, uncontrolled arrhythmias, etc.); 2) active infection or acute medical illness; 3) hemodynamic instability (systolic blood pressure < 90 mmHg or mean arterial pressure < 60 mmHg); 4) labile glycemic control (extreme swings in blood glucose levels, causing hyperglycemia or hypoglycemia); 5) inability to exercise (e.g. lower extremity amputation with no prosthesis); 6) severe musculoskeletal pain at rest or with minimal activity; 7) inability to sit, stand or walk unassisted (walking device such as cane or walker allowed); or, 8) shortness of breath at rest or with activities of daily living (NYHA Class IV).

### Trial Procedures

Before starting the study, some educational and motivational posters were installed in the dialysis center, so that all patients became familiar with the benefits of exercise and especially intradialytic exercise. Then principal investigator described the side-effects of inactivity and sedentary lifestyle to all patients. Specifically, the PI encouraged all patients to be active and provided information regarding potential benefits of intradialytic exercise. After providing written informed consent, eligible patients received a baseline assessment. Data were collected on demographic characteristics (age, sex, and time on hemodialysis), primary cause of kidney failure, and comorbidities (atherosclerotic heart disease, congestive heart failure, cerebrovascular accident/transient ischemic attack, peripheral vascular disease, dysrhythmia, and other cardiac diseases, chronic obstructive pulmonary disease, gastrointestinal bleeding, liver disease, cancer, and diabetes). Comorbidities were quantified using Charlson comorbidity index (CCI) established for dialysis patients, which included the underlying cause of kidney failure, as well as 11 comorbidities [14].

Participants were then randomized in a 1:1 ratio to either the intervention group or control group. The randomization sequence was generated by a study biostatistician who was not otherwise involved in the study using a computer-generated random schedule (using Stata 16, Stata Crop, College Station, Tx). Allocation concealment was safeguarded through the use of sequentially numbered, sealed, opaque envelopes by a specified staff member who was not involved in the study.

### Intervention

Subjects in the intervention group performed concurrent intradialytic exercise during the 2nd hour of dialysis (60-minute exercise sessions three times a week) for 6 months. The intervention was a combination of aerobic and resistance exercises. Workout time at the beginning was 30 minutes and gradually increased to 60 minutes. Each workout session included 5 minutes of warm-up, aerobic exercises, resistance exercises and finally 10 minutes of stretching exercises to cool down. The fistula arm was kept stationary thoroughly the exercise session, with necessary precautions taken into consideration. Also, the exercise protocol was not performed on the arm with AV fistula.

Exercises were individualized in a way that matched the level of physical fitness of participants (See Additional File 1). Aerobic exercises consisted of continuously performed specified movements, such as moving legs back and forth, shoulder abduction and adduction (hand without fistula), flexing and extending the knee, internally and externally rotating the leg, and abducting and adducting the leg, in time with a played beat.

The rhythm of continuous movements was adjusted by the beats per minute of the music. This meant that participants had to coordinate the movements of their arms and legs with the beats per minute of the song being played to them. In this way, the speed and intensity of aerobic exercise was controlled by the rhythm. Resistance training was performed in a semi-recumbent position and included exercises for the upper and lower limbs as well as core strength exercises using body weight, weight cuffs, dumbbells, and elastic bands of varying intensity. Chest press, shoulder press, triceps extension, straight arm shoulder flexion, shoulder horizontal abduction, seated row, supine grip, prone grip, neutral grip, bicep curl, leg abduction, plantarflexion, dorsiflexion, straight-leg/bent knee raises, knee extension, and knee flexion were all part of the resistance training program.

All of the exercises were prescribed by an exercise physiologist who also monitored the exercise sessions and helped patients with any questions they had. At the end of each session, the exercise physiologist reviewed the adherence checklists. If a person did not attend an exercise session, a counseling session was held in the presence of a nephrologist and the exercise physiologist. The reason for the individual’s non-participation was investigated and the positive effects of exercise was explained through motivational statements. When possible, patients were reminded of previous benefits of exercise they had experienced as an incentive for them to increase adherence with the training.

Participants in the control group did not undertake any specific physical activity during dialysis. All other pharmacological, dialysis, dietary and management protocols were identical for participants in both groups. Despite differences in some hematological parameters at baseline, different medications were not used for intervention group. All participants received normal bicarbonate hemodialysis, which was carried out three times a week for an average of 4 hours. Volumetric ultrafiltration control was available on all machines. The standard dialysate flow rate was 500 mL/min and blood flow rates were prescribed according to the participant’s needs. Automated methods were applied to perform dialyzer reuse uniformly.

### Blood sampling

Baseline blood samples were collected one day before the start of the exercise session. Exercise began at the mid-week dialysis session. After the end of the 36th session (end of the third month) and after the end of the 72nd session (end of the sixth month), subsequent blood samples were collected the day before the midweek dialysis session. The control group was assessed at the same time points. On a nondialysis day, blood samples were taken from the arterial needle after at least an 8-hour fast. Approximately 30 milliliters of blood were collected and centrifuged for 15 minutes at 20 °C and 2500 g. Plasma was next pipetted into cryotubes and stored at -80 °C in a freezer that was electronically monitored. All samples were measured in duplicate, in line with the manufacturers’ suggested protocol, and within the manufacturer’s specified range of acceptable variation and sensitivity.

### Outcomes

The primary outcome measures included changes in serum calcium (mg/dL), serum phosphorous (mEq/L) and parathyroid hormone (pg/mL) over time. Rate of changes of alkaline phosphatase (U/L) and calcium-phosphorous product (mg^2^ /dL^2^) were the secondaries. These outcomes were evaluated at baseline, 3 months and 6 months.

Safety outcomes included all serious adverse events and adverse events.

### Adherence

Intervention adherence was defined as the number of sessions performed divided by the number of sessions offered, multiplied by 100.

### Blinding

Due to the nature of the intervention, it was not feasible to blind participants or study staff. However, outcome assessors and data analysts were blinded to participants’ treatment allocations.

### Sample size

The sample size was calculated in accordance with a previous study [23], by NCSS PASS 16.0 software. The model was established according to the Repeated Measure ANOVA (bilateral side) for the PTH as main variable, with α=0.05 and power 1 – β = 0.8.

Assuming an effect size of 0.45 for reduction of PTH and a drop-out rate of 20%, 44 participants (22 per group) were required to provide 80% power.

### Statistical analysis

Data are presented as frequency (percentage), mean ± standard deviation, or median and interquartile range, depending on data type and distribution. A detailed statistical analysis plan was prepared and completed prior to database lock. Primary and secondary outcomes were evaluated using Repeated Measure ANOVA and the Friedman test. Statistical analyses were performed using IBM SPSS software 25. P values less than 0.05 were considered statistically significant.

## Results

Overall, 58 patients were assessed for eligibility, of whom 44 were consented and randomized. The corresponding flowchart is presented in Figure 1.

**Figure 1.**
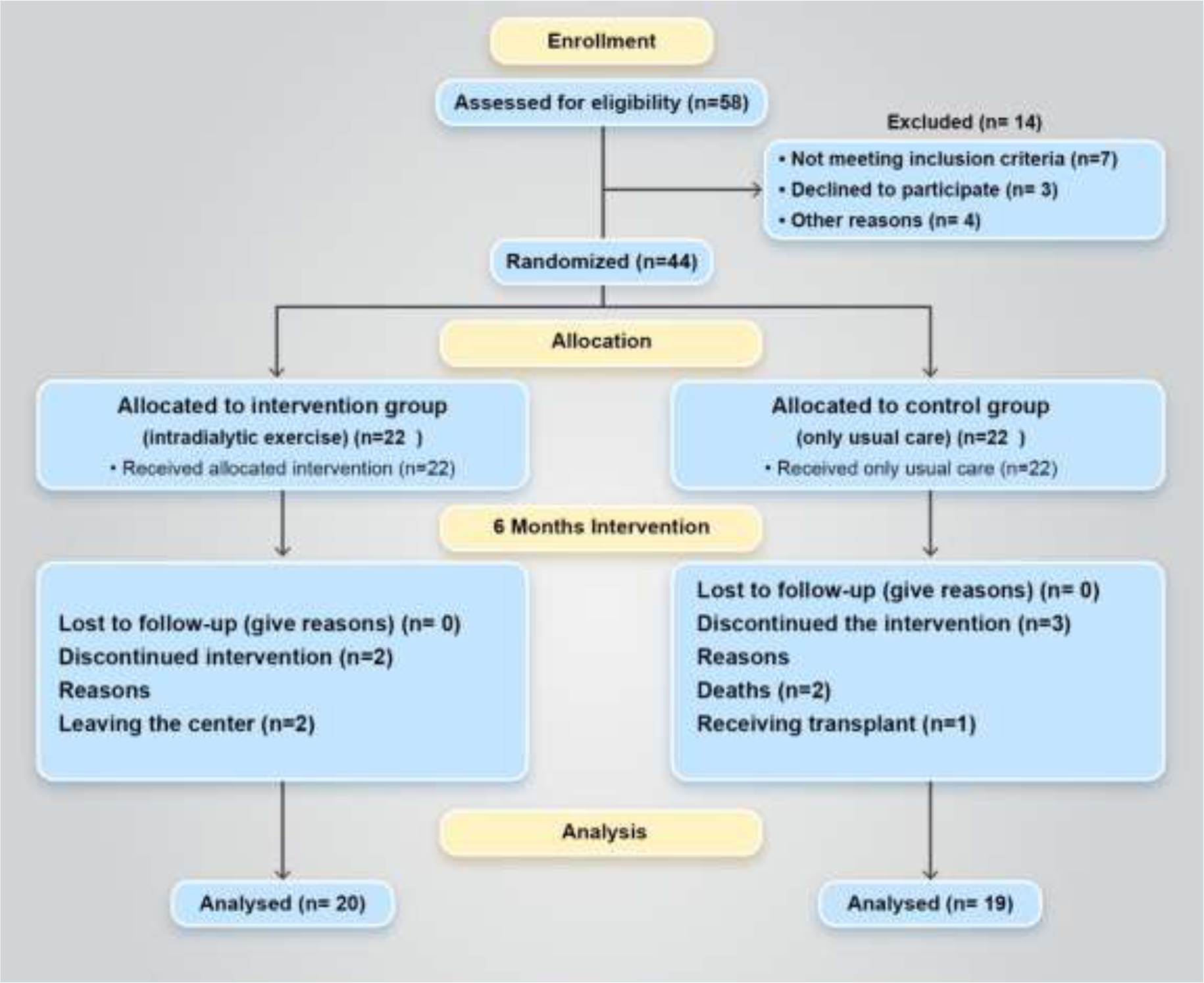
Participant flow during the study.

During the 6-month intervention period, 2 participants in the intervention group and 3 participants in the control group dropped out of the study.

### Baseline characteristics

Baseline characteristics were balanced between the assigned treatment groups (Table 1).

**Table 1.**
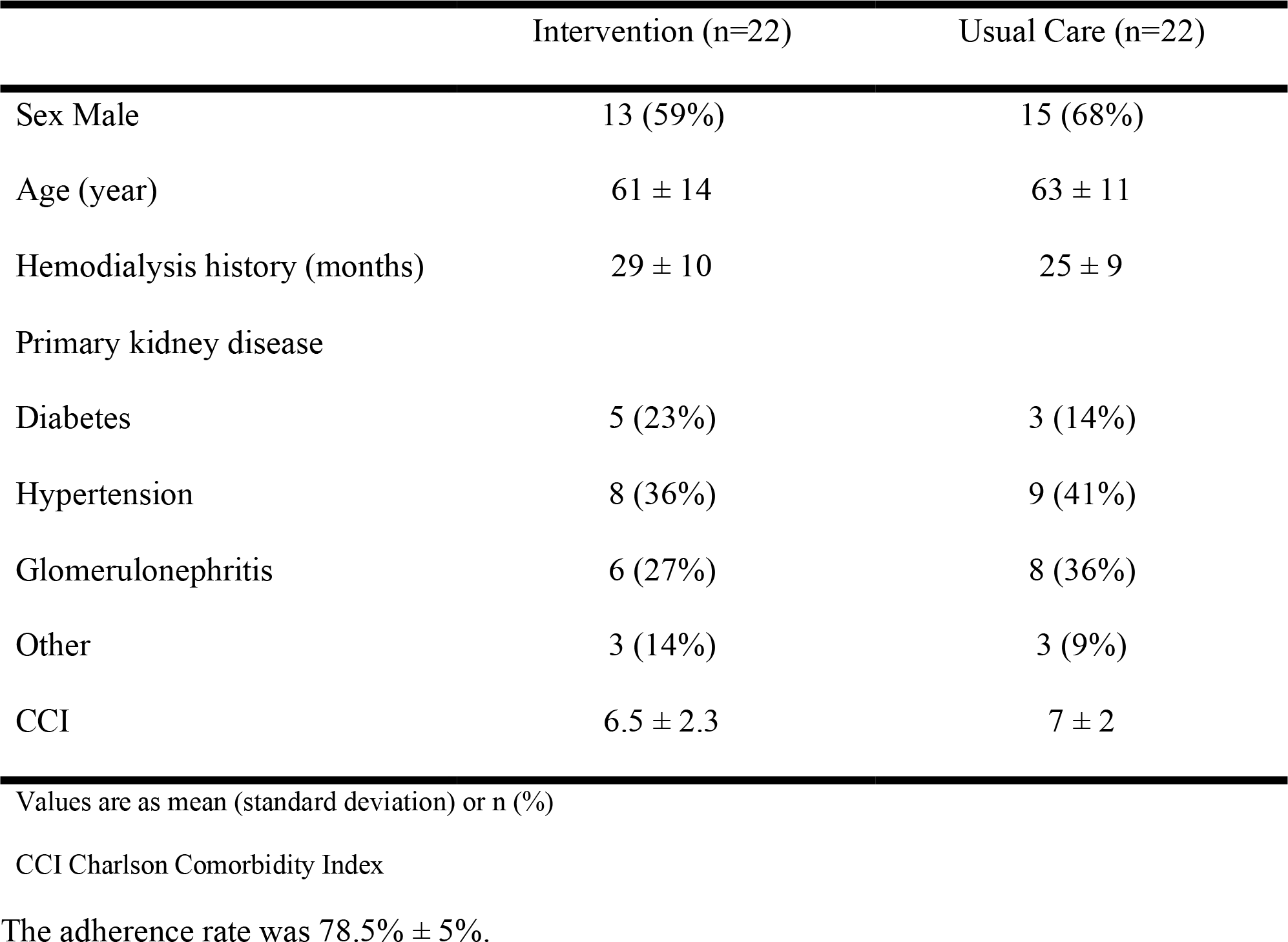
Baseline characteristics of patients

### Outcome analysis

Significant between-group changes during the 6-month intervention period were observed in all primary and secondary outcomes (Table 3) between the intervention and control groups. Specifically, serum calcium tended to increase in the intervention group, but remained relatively stable in the control group. In contrast, serum parathyroid hormone, phosphorus, calcium-phosphorous product and alkaline phosphatase levels significantly decreased in the intervention group but remained relatively stable in the control group (Table 3).

**Table 3.** Change of primary and secondary outcomes and the results of longitudinal analysis

|  | Intervention |  |  | Usual care |  |  | Partial<br>Eta<br>Square | Group<br>×Time |
| --- | --- | --- | --- | --- | --- | --- | --- | --- |
|  | Baseline<br>(n=22) | 3 months<br>(n=22) | 6 months<br>(n=20) | Baseline<br>(n=22) | 3 months<br>(n=21) | 6 months<br>(n=19) |  |  |
| Ca (mg/dl) | 8.1 ± 0.8 | 8.7 ± 0.6 | 9.2 ± 0.6 | 8.4 ± 0.7 | 8.6 ± 0.7 | 8.5 ± 0.6 | 0.5 | <0.001** |
| P (meq/lit) | 5.8 ± 1.4 | 5.1 ± 1.1 | 4.1 ± 0.5 | 5.4 ± 1.3 | 5.9 ± 1.2 | 5.3 ± 0.9 | 0.4 | 0.002** |
| PTH (pg/ml) | 441<br>(364 - 597) | 290<br>(224 - 391) | 245 †<br>(173 - 286) | 270<br>(180 - 418) | 342<br>(245 - 476) | 312<br>(243 - 407) | 0.8 | NA |
| Ca × P (mg<sup>2</sup>/dL<sup>2</sup>) | 45.5 ± 4.6 | 43 ± 5.1 | 35.9 ± 4.1 | 43.8 ± 5.5 | 50.5 ± 4.9 | 44.2 ± 5 | 0.5 | 0.04* |
| ALP (U/L) | 195.5 ± 8.6 | 148 ± 5.5 | 125.4 ± 7.9 | 203.5 ± 7.8 | 221.1 ± 8.3 | 211 ± 10.5 | 0.9 | <0.001** |

Ca serum calcium, P serum phosphorous, PTH parathyroid hormone, ALP alkaline phosphatase
Values are as mean ± standard deviation or median (interquartile)
Group ×Time is an interaction from a two-way repeated-measures analysis of the effect of time from baseline to 6 months
\*p < 0.05 significant
\*\*p < 0.01 highly significant
†p < 0.01 highly significant within-group difference, analyzed by Friedman test

### Safety

No treatment-related serious adverse effects were observed during the period of the study. During the intervention period, one patient had muscle cramps after the first exercise session, but this was not serious or harmful.

## Discussion

Renal hyperparathyroidism is a common complication of chronic kidney disease characterized by elevated PTH levels secondary to derangements in the homeostasis of calcium, phosphate, and vitamin D [24].

Abnormalities in calcium, phosphorous, and PTH, hallmarks of the condition known as chronic kidney disease – mineral and bone disorder (CKD-MBD) [25], are associated with adverse outcomes in patients receiving maintenance dialysis [7,8,26]. In patients with advanced chronic kidney disease results in derangements of circulating markers of mineral metabolism and in dysregulation of skeletal and cardiovascular physiology [27-29]. Epidemiologic studies of dialysis patients provide substantial evidence that elevated PTH is associated with mortality [8,26]. However, therapies targeting abnormal CKD-MBD parameters, while improving biochemical end points [30], have failed to convincingly demonstrate reductions in “hard” end points such as all-cause and cardiovascular mortality in clinical trials [31]. The Kidney Disease Improving Global Outcomes guidelines recommend that screening and management of hyperparathyroidism be initiated for all patients with chronic kidney disease [25].

The present study demonstrated that the 6-month intradialytic concurrent exercise program was effective in improving PTH levels, reducing serum phosphorous and improving serum calcium and calcium-phosphorous product as well. However, contradictory results have been reported by some other studies.

Previous work showed that exercise decreased plasma PTH levels in CKD patients [32]. Research on effect of exercise on PTH is limited, but it is affected by physical activity. Its levels can vary after physical exercise depending on the duration and intensity of the activity [33]. Moreover, resistance to PTH action can occur with progression of renal disease [34].

In the present study, phosphorus levels in the intervention group were significantly lower than the baseline. The effect of exercise on phosphorus level was in line with findings of the several previous studies [35,36]. Borzou et al. also reported that increasing the blood flow to 250 mL/min and then to 300 mL/ min can be effective in clearance of phosphorus from the blood [36].

Reviewing the limited available research on the effects of exercise in reducing phosphorus levels showed that although exercise decreased the level of phosphorus, the significant effects and changes could be observed in long-term and perhaps more intense exercise might be required for some studies [35,37,38].

This study revealed that serum calcium levels showed significant change in the intervention group but Saralhab et al. like some other researchers failed to retrieve any influence of exercise on serum calcium [24,34,39]. Perhaps studies with longer time frame and differences in type and frequency of exercise are needed.

Cardoso et al. in their systemic review found evidence supporting a positive relation between physical activity and bone outcomes in patients with CKD [40]. Studies show that exercise acts as a regulator of homeostasis, alters the levels of circulatory mediators and hormones, and increases the demand for skeletal muscle and other vital organs for energy substrates. Exercise also activates bone and mineral metabolism, especially calcium and phosphate, both of which are essential for muscle contraction, neuromuscular signaling, adenosine triphosphate biosynthesis, and other energy substrates. Due to the fact that PHT is involved in the regulation of calcium and phosphate homeostasis, chronic exercise may help in limiting PTH secretion exercise activity by changing the level of calcium and phosphate in the bloodstream. On the other hand, PTH reacts directly to exercise and exercise myokines. [33].

Alkaline phosphatase is an enzyme measurable in most body fluids and usually originates from the liver or bone. In CKD patients without liver disease, ALP is a surrogate of high turnover bone disease and is used to monitor the metabolic bone disease associated with renal insufficiency [41-43]. Furthermore, elevated ALP may be causally involved in the cardiovascular calcification of CKD [44-46], making it a potentially important independent risk factor. Higher ALP has been shown to be associated with mortality and coronary artery calcification in CKD 5D [43] and in patients without CKD [47]. In CKD, ALP is more reliable than many other biochemical bone markers because serum concentrations of ALP are not influenced by the glomerular filtration rate [47]. An association exists between higher serum ALP and worse dual-energy X-ray absorptiometry-assessed BMD in HD patients [48].

ALP levels significantly decreased in exercise group, but controls showed no similar change.

Previous observational studies on dialysis patients revealed a relationship between physical activity serum levels of ALP due to the fact that energy expenditure has a strong relationship to the total BMD [49,50].

The study by Gomes et al. and the other one by Elshinnawy et al. showed there was significant increase in ALP levels following exercise in patients with CKD [24,51].

One of the reasons for the contradiction in the process of ALP changes with the previous studies is that in those studies, the ALP level of the patients was within the normal range before the intervention, and the intervention has somehow improved the condition. However, in the current study, the ALP level of the patients was much higher than normal, and exercise has been able to have a significant positive effect on improving the patients’ condition by reducing the ALP level.

As levels of bone specific ALP increase during weight-bearing exercise [52], but return to baseline within 20-minutes following exercise, lower resting levels may be a consequence of the transient response to weight bearing exercises [53].

Based on Mechanostat theory which was developed by Harold frost, the local deformation from the mechanical loading stimulates bone cells and results in bone adaptation (it means that bone tissues accommodate to the stress applied on it) [54, 55]. Without any doubt bones are subjected to two sources of load including ground reaction force and muscular force [54].

High proinflammatory cytokine levels may contribute to bone loss and fracture in patients with CKD [51]. In addition, inflammatory cytokines are potent activators of receptor activator of nuclear factor kappa B ligand / receptor activator of nuclear factor kappa B (RANKL/RANK)-activated osteoclast genesis [55]. Exercise may inactivate the RANKL/RANK pathway by ameliorating inflammation and preventing bone loss in HD patients.

A major strength of the present study was the fact that all exercises were tailored according to each individual’s functional status within a pre-specified structure. There was also a high participation rate, such that included HD patients exhibited considerable diversity with respect to demographic characteristics and associated comorbidities. In this study also, is that, at the end of the study, the level of calcium, phosphorus and parathyroid hormone in the intervention group reached the level recommended by KDOQI guidelines [25].

This discrepancy between the results of the present study and the findings of the above-mentioned studies can be attributed to differences in participants’ age and sex or duration, intensity, and type of exercise program. The length of the other studies is shorter. Moreover, the intradialytic intervention was performed as concurrent exercise which was different from other studies, performing only resistance or aerobic exercise.

Balanced against these strengths, the study also had a number of limitations. Not measuring the effect of exercise on bone mass and cardiovascular indexes are limitations of this study. Indeed, not measuring bone mineral density in this work is a limitation as bone strength depends on bone quality and quantity. Future work should focus on observing the effect of different types of exercise on bone mineral disorders and all-cause mortality in HD patients.

## Conclusion

Intradialytic exercise performed for at least 60 minutes during thrice weekly dialysis sessions improves bone mineral metabolism in adult patients receiving HD and perhaps decrease all-cause mortality in these patients. Further studies should focus on observing the effect of different types of exercise on bone mineral disorders and all-cause mortality in HD patients.

## Data Availability

All data produced in the present study are available upon reasonable request to the authors.

## Declaration

### Ethics approval and consent to participate

The study protocol and informed consent were approved by Iran National Committee for Ethics in Biomedical Research (approval number IR.IAU.KHUISF.REC.1400.088) and was conducted in accordance with principles of the Declaration of Helsinki. All patients provided written informed consent prior to enrollment.

## Consent for publication

Not applicable

## Authors’ Contribution

MAT: conceptualizing the study, project leader of the study, conducting the study, writing of the manuscript, supervising of the analysis, interpreting the results, approval of the manuscript, KW: conceptualizing of the study, co-writing of the manuscript, supervising of the manuscript, approval of the manuscript, NS: project leader of the study, analyzing the data, interpreting the results, co-writing of the manuscript, approval of the manuscript, SN: conceptualizing the study, conducting the study, co-writing of the manuscript, supervising of the manuscript, approval of the manuscript, MS: conceptualizing the study, conducting the study, co-writing of the manuscript, supervising of the manuscript, approval of the manuscript, ZR: conceptualizing the study, conducting the study, co-writing of the manuscript, supervising of the manuscript, approval of the manuscript, SA: conceptualizing the study, conducting the study, co-writing of the manuscript, approval of the manuscript. Each author contributed with important intellectual content during the manuscript drafting or revision and accepts accountability for the overall work by ensuring that questions pertaining to the accuracy or integrity of any portion of the work are appropriately investigated and resolved. All authors read and approved the final manuscript.

## Acknowledgement

The authors would like to express their heart-felt gratitude to all the investigators for their contribution to the trial, especially Dr. Hugo Corrêa1 and Dr. João and all the staff of the dialysis center for their efforts and patience in helping to maintain the standards of the research, as well as all patients involved in this study. Also, the authors would like to thank Pardis Specialized Wellness Institute and all the personnel of the institute for their support throughout the study. The authors would also like to thank the opportunity that medRxiv offered them to deposit the preprint version.

## References

1. Ortiz A, Covic A, Fliser D, Fouque D, Goldsmith D, Kanbay M, et al. Epidemiology, contributors to, and clinical trials of mortality risk in chronic kidney failure. Lancet. 2014;383(9931):1831–43

2. United States Renal Data System. Epidemiology of kidney disease in the United States. National Institutes of Health, National Institute of Diabetes and Digestive and Kidney Diseases, Bethesda, MD, 2018. Available at https://www.usrds.org/2018/view/Default.aspx.

3. Deligiannis A, D’Alessandro C, Cupisti A. Exercise training in dialysis patients: impact on cardiovascular and skeletal muscle health. Clin Kidney J. 2021 Jan 11;14(Suppl 2):ii25–ii33. doi: 10.1093/ckj/sfaa273. PMID: 33981417; PMCID: PMC8101623.

4. SONG Initiative. The SONG Handbook. Version 1.0 June 2017, Sydney,Australia Available at songinitiative.org/reports-and-publications/.

5. Manera KE, Tong A, Craig JC, Shen J, Jesudason S, Cho Y, et al. An international Delphi survey helped develop consensus-based core outcome domains for trials in peritoneal dialysis. Kidney Int. 2019;96(3):699–710.

6. D. S. March, M. P. M. Graham-Brown, C. M. Stover, N. C. Bishop, and J. O. Burton, “Intestinal Barrier sturbances in Haemodialysis Patients: Mechanisms, Consequences, and Therapeutic Options,” BioMed Research International, vol. \v2017, Article ID 5765417, 2017.

7. Block GA, Hulbert-Shearon TE, Levin NW et al. Association of serum phosphorus and calcium phosphate product with mortality risk inchronic hemodialysis patients: a national study. Am J Kidney Dis 1998; 31: 607–617.

8. Block GA, Klassen PS, Lazarus JM et al. Mineral metabolism, mortality, and morbidity in maintenance hemodialysis. J Am Soc Nephrol 2004; 15: 2208–2218.

9. Ganesh SK, Stack AG, Levin NW et al. Association of elevated serum PO(4), Ca PO(4) product, and parathyroid hormone with cardiac mortality risk in chronic hemodialysis patients. J Am Soc Nephrol 2001; 12: 2131–213

10. Stevens LADO, Cardew S, Cameron EC, Levin A. Calcium, phosphate, and parathyroid hormone levels in combination and as a function of dialysis duration predict mortality: evidence for the complexity of the association between mineral metabolism and utcomes. J Am Soc Nephrol 2004; 15: 770.

11. Slinin Y, Foley RN, Collins AJ. Calcium, phosphorus, parathyroid hormone, and cardiovascular disease in hemodialysis patients: the USRDS waves 1, 3, and 4 study. J Am Soc Nephrol 2005; 16: 1788–1793.

12. Albuquerque RFC, Carbonara CEM, Martin RCT, Reis LM, Nascimento Jr CP, Arap SS, et al. Parathyroidectomy in patients with chronic kidney disease: Impacts of different techniques on the biochemical and clinical evolution of secondary hyperparathyroidism. Surgery. 2018;163(2):381–7

13. Campos SR, Gusmão MHL, Fortes A, Sampaio LR, Barreto JM. Estado nutricional e ingestão Estado nutricional e ingestão alimentar de pacientes em diálise peritoneal contínua com e sem hiperparatireoidismo secundário. J Bras Nefrol. 2012;34(2):170–7

14. Coelho DM, Castro ADM, Tavares HA, Abreu PBC, Glória RR, Duarte MH, et al. Efeitos de um Programa de Exercícios Físicos no Condicionamento de Pacientes em Hemodiálise. J Bras Nefrol. 2006;28(3):121–7.

15. Duarte J, Medeiros RF, Di Pietro T, Lopes TM. Alterações de volumes e capacidades pulmonares pré e pós-hemodiálise em insuficiência renal crônica TT Changes in lung volume and capacity before and after hemodialysis in chronic renal failure. J Heal Sci Inst. 2011;29(1):70–2

16. Fassbinder TRC, Winkelmann ER, Schneider J, Wendland J, Oliveira OB. Functional Capacity and Quality of Life in Patients with Chronic Kidney Disease In Pre-Dialytic Treatment and on Hemodialysis - A Cross sectional study. J Bras Nefrol. 2015;37(1):47–54

17. Maïmoun, L., Lumbroso, S., Manetta, J., Paris, F., Leroux, J.L. and Sultan, C. (2003) Testosterone is significantly reduced in endurance athletes without impact on bone mineral density. Hormone Research 59, 285–292

18. Glastre, C., Braillon, P., David, L., Cochat, P., Meunier, P.J. and Delmas P.D. (1990) Measurement of bone mineral content of the lumbar spine by dual energy x-ray absorptiometry in normal children: correlations with growth parameters. The Journal of Clinical Endocrinology and Metabolism 70, 1330–1333.

19. Thorsen, K., Kristoffersson, A., Hultdin, J. and Lorentzon, R. (1997) Effects of moderate endurance exercise on calcium, parathyroid hormone, and markers of bone metabolism in young women. Calcified Tissue International 60, 16–20.

20. Brahm, H., Piehl-Aulin, K. and Ljunghall, S. (1997a) Bone metabolism during exercise and recovery: the influence of plasma volume and physical fitness. Calcified Tissue International 61, 192–198.

21. Ljunghall, S., Joborn, H., Roxin, L.E., Skarfors, E.T., Wide, L.E. and Lithell, H.O. (1988) Increase in serum parathyroid hormone levels after prolonged physical exercise. Medicine and Science in Sports Exercise 20, 122–125

22. Zerath, E., Holy, X., Douce, P., Guezennec, C.Y. and Chatard, J.C. (1997) Effect of endurance training on postexercise parathyroid hormone levels in elderly men. Medicine and Science in Sports and Exercise 29, 1139–4115.

23. Elshinnawy, H.A., Mohamed, A.M.B.B., Farrag, D.A.B. et al. Effect of intradialytic exercise on bone profile in hemodialysis patients. Egypt Rheumatol Rehabil 48, 24 (2021).

24. Yuen NK, Ananthakrishnan S, Campbell MJ. Hyperparathyroidism of Renal Disease. Perm J. 2016 Summer;20(3):15–127. doi: 10.7812/TPP/15-127. Epub 2016 Jul 22.

25. Kidney Disease: Improving Global Outcomes (KDIGO) CKD-MBD Work Group (2017) KDIGO 2017 Clinical practice guideline update for the diagnosis,evaluation, prevention, and treatment of chronic kidney disease-mineral and bone disorder (CKD-MBD). Kidney Int Suppl 7:1–59. 2009;76(Suppl 113):S1–S130.

26. Tentori F, Wang M, Bieber BA, et al. Recent changes in therapeutic approaches and association with outcomes among patients with secondary hyperparathyroidism on chronic hemodialysis: the DOPPS study. Clin J Am Soc Nephrol. 2015;10(1):98–109.

27. Nickolas TL, Leonard MB, Shane E. Chronic kidney disease and bone fracture: a growing concern. Kidney Int. 2008;74(6):721–731.

28. Hruska KA, Choi ET, Memon I, Davis TK, Mathew S. Cardiovascular risk in chronic kidney disease (CKD): the CKD-mineral bone disorder (CKD-MBD) Pediatr Nephrol. 2010;25(4):769–778.

29. Faul C, Amaral AP, Oskouei B, et al. FGF23 induces left ventricular hypertrophy. J Clin Invest. 2011;121(11):4393–4408.

30. Moe SM, Chertow GM, Coburn JW, et al. Achieving NKF-K/DOQI bone metabolism and disease treatment goals with cinacalcet HCl. Kidney Int. 2005;67(2):760–771.

31. Chertow GM, Block GA, Correa-Rotter R, et al. Effect of cinacalcet on cardiovascular disease in patients undergoing dialysis. N Engl J Med. 2012;367(26):2482–2494.

32. Bouassida A, Latiri I, Bouassida S, Zalleg D, Zaouali M, Feki Y, Gharbi N, Zbidi A, Tabka Z (2006) Parathyroid hormone and physical exercise: a brief review. J Sports Sci Med 5(3):367–374

33. Lombardi G, Ziemann E, Banfi G, Corbetta S (2020) Physical activity-dependent regulation of parathyroid hormone and calcium-phosphorous metabolism. Int J Mol Sci 21(15):5388.

34. Makhlough A, Ilali E, Mohseni R, Shahmohammadi S. Effect of intradialytic aerobic exercise on serum electrolytes levels in he-modialysis patients. Iran J Kidney Dis. 2012;6(2):119–23.

35. Vaithilingam I, Polkinghorne KR, Atkins RC, Kerr PG. Time and exercise improve phosphate removal in hemodialysis patients. Am J Kidney Dis. 2004;43(1):85–9.

36. Borzou SR, Gholyaf M, Zandiha M, Amini R, Goodarzi MT, Torkaman B. The effect of increasing blood flow rate on dialysis adequacy in hemodialysis patients. Saudi J Kidney Dis Transpl. 2009;20(4):639–42.

37. Asadi Noghabi A, Bassampour SH, Zolfaghari M. Critical care nursing ICU, CCU, dialysis. Salemi publication; 2th ed. 2007; p. 448–53.

38. Cappy CS, Jablonka J, Schroeder ET. The effects of exercise during hemodialysis on physical performance and nutrition assessment. J Ren Nutr. 1999;9:63–70.

39. Salhab N, Alrukhaimi M, Kooman J, Fiaccadori E, Aljubori H, Rizk R, Karavetian M (2019) Effect of intradialytic exercise on hyperphosphatemia and malnutrition. Nutrients 11(10):2464.

40. Cardoso, D.F., Marques, E.A., Leal, D.V. et al. Impact of physical activity and exercise on bone health in patients with chronic kidney disease: a systematic review of observational and experimental studies. BMC Nephrol 21, 334 (2020).

41. Magnusson P, Sharp CA, Magnusson M, Risteli J, Davie MW, Larsson L: Effect of chronic renal failure on bone turnover and bone alkaline phosphatase isoforms. Kidney Int 60: 257–265, 2001

42. Torres PU. Bone alkaline phosphatase isoforms in chronic renal failure. Kidney Int. 2002;61:1178–1179.

43. Regidor DL, Kovesdy CP, Mehrotra R, et al. Serum alkaline phosphatase predicts mortality among maintenance hemodialysis patients. J Am Soc Nephrol. 2008;19:2193–2203.

44. Lomashvili KA, Garg P, Narisawa S, Millan JL, O’Neill WC. Upregulation of alkaline phosphatase and pyrophosphate hydrolysis: potential mechanism for uremic vascular calcification. Kidney Int. 2008;73:1024–1030.

45. O’Neill WC. Pyrophosphate, alkaline phosphatase, and vascular calcification. Circ Res. 2006;99:e2.

46. Schoppet M, Shanahan CM. Role for alkaline phosphatase as an inducer of vascular calcification in renal failure?. Kidney Int. 2008;73:989–991.

47. Tonelli M, Curhan G, Pfeffer M, et al. Relation between alkaline phosphatase, serum phosphate, and all-cause or cardiovascular mortality. Circulation. 2009;120:1784–1792.

48. Park JC, Kovesdy CP, Duong U, Streja E, Rambod M, Nissenson AR, Sprague SM, Kalantar-Zadeh K. Association of serum alkaline phosphatase and bone mineral density in maintenance hemodialysis patients. Hemodial Int. 2010 Apr;14(2):182–92. doi: 10.1111/j.1542-4758.2009.00430.x. Epub 2010 Mar 24.

49. Huang G-S, Chu T-S, Lou M-F, Hwang S-L, Yang R-S. Factors associated with low bone mass in the hemodialysis patients - a cross-sectional correlation study. BMC Musculoskelet Disord. 2009;10:60.

50. Ota S, Takahashi K, Taniai K, Makino H. Bone metabolism and daily physical activity in women undergoing hemodialysis. Nihon Jinzo Gakkai Shi. 1997;39(4):441–6.

51. Gomes TS, Aioke DT, Baria FG, Graciolli FG, Moyses RMA, Cuppari L (2017) Effect of aerobic exercise on markers of bone metabolism of overweight and obese patients with chronic kidney disease. J Ren Nutr 27(5):364–371.

52. Rudberg A, Magnusson P, Larsson L, Joborn H. Serum isoforms of bone alkaline phosphatase increase during physical exercise in women. Calcif Tissue Int. 2000;66: 342–347. pmid:10773103

53. Fragala MS, Bi C, Chaump M, Kaufman HW, Kroll MH (2017) Associations of aerobic and strength exercise with clinical laboratory test values. PLOS ONE 12(10): e0180840.

54. Frost HM. The mechanostat: a proposed pathogenic mechanism of osteoporoses and the bone mass effects of mechanical and nonmechanical agents. Bone Miner. 1987;2(2):73–85.

55. Frost HM. Bone’s mechanostat: a 2003 update. Anat Rec A Discov Mol Cell Evol Biol. 2003;275(2):1081–1101.

56. Liao M-T, Liu W-C, Lin F-H, Huang C-F, Chen S-Y, Liu C-C, et al. Intradialytic aerobic cycling exercise alleviates inflammation and improves endothelial progenitor cell count and bone density in hemodialysis patients. Medicine. 2016;95(27):e4134.

